# Medication history, not clinical profiles, predicts antihypertensive treatment persistence across health systems

**DOI:** 10.64898/2026.11.02.26367145

**Authors:** Shupeng Wang, Dongdong Lu, Tao Tao

**Affiliations:** Department of Pharmacy, South China Hospital of Shenzhen University, Shenzhen, China; Department of Experimental Research, South China Hospital of Shenzhen University, Shenzhen, China; Department of Nutritional Physiology, Institute of Nutritional Sciences, Friedrich Schiller University Jena, Jena, Germany; Shenzhen Luohu People’s Hospital; The Third Affiliated Hospital (The Affiliated Luohu Hospital) of Shenzhen University, Shenzhen, China

**Author notes:** Shupeng Wang and Dongdong Lu contributed equally to this work. Correspondence: Shupeng Wang; Tao Tao.

**Keywords:** antihypertensive persistence, medication adherence, machine learning, external validation, model transportability, CHARLS, SHARE, HRS

## Abstract

**Background:** Hypertension control remains poor worldwide, in large part because patients discontinue antihypertensive treatment. Machine-learning (ML) prediction models for medication persistence are typically single-country and cross-sectional, rarely compared with simple clinical information, and lack external validation across health systems.

**Methods:** We harmonized three prospective aging cohorts — the China Health and Retirement Longitudinal Study (CHARLS), the Survey of Health, Ageing and Retirement in Europe (SHARE), and the US Health and Retirement Study (HRS) — including adults aged ≥50 years with doctor-diagnosed hypertension at baseline and known medication status at follow-up. We used baseline plus two prior waves to predict non-use of any antihypertensive medication at the next wave (treatment non- persistence), with 21 harmonized cross-sectional features, 11 longitudinal history features (including the prior non-use ratio), and a 3-feature parsimonious medication-history score (baseline non-use, prior non-use ratio, prior discontinuation). Models were L2-regularized logistic regression and random forest, evaluated by 5-fold × 5-repeat nested cross-validation and by direct transport of a CHARLS-trained model to SHARE and HRS without retraining. Three simple rule-based comparators were prespecified: baseline carry-forward (T0), prior non-use count (T1), and the 3-feature score (T2). Non-inferiority of T2 versus the full 32-feature logistic model was tested at δ=0.03 AUC. Country-level external validation across 18 SHARE countries used random-effects meta-analysis and inverse-variance meta-regression on World Bank indicators.

**Results:** The analytic samples comprised 4,532 (CHARLS), 26,128 (SHARE) and 9,495 (HRS) participants, with non-persistence of 22.8%, 19.1% and 12.6%. Within each cohort, adding medication history raised AUC from 0.59–0.67 to 0.78–0.85 (ΔAUC 0.17–0.20; all 95% CIs >0). On direct transport, history models retained AUC 0.73–0.76 in SHARE and 0.83–0.85 in HRS, whereas cross- sectional models collapsed to near chance in SHARE (AUC 0.55–0.57). The baseline carry-forward rule alone achieved AUC 0.70–0.80; the 3-feature score achieved 0.75–0.83 and was non-inferior to the full logistic model (ΔAUC ≤0.027; upper CI bounds <0.03) in all three within-cohort comparisons and both transport targets. Across 18 European countries, cross-sectional models pooled at 0.55 (I²=30%) while history models pooled at 0.74–0.76 (I²=82–88%); transported AUC increased with national wealth (r=0.58, P=0.011), and calibration slope declined with non-persistence burden (r=−0.69, P=0.002) and out-of-pocket expenditure share (r=−0.51, P=0.032).

**Conclusions:** A parsimonious medication-history core transfers across health systems, whereas cross- sectional clinical profiles do not. Complex ML adds little beyond a one-question or three-variable medication-history rule. Discrimination is strongest in high-resource, low-burden systems, but probability calibration degrades where non-persistence is common, requiring local recalibration before individual-level use.

## 1. Background

Hypertension is the leading preventable cause of cardiovascular morbidity and mortality worldwide, affecting more than one billion adults. Although blood-pressure lowering substantially reduces stroke and cardiac events, treatment control rates remain low in every income setting. A dominant reason is failure to persist with antihypertensive treatment: many patients never initiate therapy, and a substantial proportion discontinue within the first year. Identifying individuals at high risk of non-persistence is central to improving hypertension outcomes, particularly in aging populations in whom chronic disease management is most intensive [1–6].

Machine learning is increasingly used to predict medication adherence and persistence, because it can accommodate nonlinear and interacting relationships among sociodemographic, clinical, psychological and social determinants [7,8]. However, the published literature has three recurrent limitations. First, most models are developed and tested within a single country or institution, often with a single train– test split, so their transportability to other health systems is unknown [9–12]. Second, many models rely on cross-sectional predictors measured at the same time as the outcome, or on outcome definitions that conflate current treatment status with adherence, which can inflate apparent discrimination. Third, models are rarely benchmarked against the simplest clinically available information — such as whether the patient is currently taking the medication or has previously stopped it — so it remains unclear how much complex ML adds beyond routine questions [12–14].

Longitudinal medication history has strong face validity for predicting persistence: the strongest predictor of future behavior is past behavior, a principle long recognized in adherence research [15]. Electronic prescribing and pharmacy refill records make such history increasingly available in routine care. Yet prior studies have generally used past refill behavior only in the context of concurrent adherence measurement, rather than as a prospective predictor of treatment persistence across multiple health systems [15–17].

In this study we address these gaps using three large, harmonized prospective cohorts of adults with doctor-diagnosed hypertension — CHARLS (China), SHARE (Europe) and HRS (USA). We asked four questions: (i) How much does adding longitudinal medication history improve prediction of future antihypertensive non-persistence within cohorts, over and above cross-sectional clinical profiles? (ii) Does a CHARLS-trained model transport to SHARE and HRS without retraining? (iii) How much of the full model’s discrimination is captured by simple rule-based comparators, including a 3-feature parsimonious medication-history score, and is the score non-inferior to the full model at a prespecified margin? (iv) Across 18 European countries, where and why do history-based models work — do health- system characteristics explain country-level variation in transported discrimination and calibration?

## 2. Methods

### 2.1 Data sources

We used harmonized data from three population-representative prospective cohort studies of adults aged ≥50 years: the China Health and Retirement Longitudinal Study (CHARLS, harmonized waves 2011–2018, baseline waves up to 2015 with 3-year follow-up to 2018), the Survey of Health, Ageing and Retirement in Europe (SHARE, harmonized waves 2011–2017, 18 countries), and the US Health and Retirement Study (HRS, harmonized waves 2012–2018) [18–22]. All variables were drawn from the Gateway to Global Aging Data harmonized files to maximize cross-cohort comparability (rxhibp medication-use variables and harmonized health measures) [22–24]. Baseline was defined as the first wave at which a participant reported doctor-diagnosed hypertension and had complete medication information; the two preceding waves (where available) contributed the longitudinal medication history. For participants entering at the first available wave of each cohort, history features were defined using baseline information only (see Supplementary Methods S1.1). Temporal external validation additionally used the most recent available waves (SHARE 2017→2020, n=2,851; HRS 2018→2020, n=211; see Section 3.6).

### 2.2 Study population and outcome

Eligible participants were aged ≥50 years, had doctor-diagnosed hypertension at baseline, and had known medication status at both baseline and the outcome wave. The outcome was treatment non- persistence: self-reported non-use of any antihypertensive medication at the follow-up wave among participants who had been prescribed or had ever used antihypertensives (positive class = 1). Follow- up intervals were three years in CHARLS and two years in SHARE and HRS. Participants with missing medication status at the outcome wave were excluded. The final analytic samples comprised 4,532 (CHARLS), 26,128 (SHARE) and 9,495 (HRS) participants.

### 2.3 Predictors and feature sets

We constructed 21 harmonized cross-sectional features (demographics, education, marital status, self- rated health, diagnosed chronic conditions, functional limitations, depressive symptoms, smoking, alcohol, BMI, recent hospitalization and doctor contact) and 11 longitudinal features derived from waves before the outcome (number of observed waves, adherent and non-adherent waves, ever-adherent, ever-non-adherent, state-change count, baseline medication status, prior non-use ratio, prior discontinuation, known diagnosis before baseline, years since diagnosis). Three feature sets were compared: cross-sectional (21 features), full (32 features = 21 + 11), and a parsimonious 3-feature medication-history score (T2): baseline non-use (base_nonadh), prior non-use ratio (nonadh_ratio = number of non-adherent waves / number of observed waves), and prior discontinuation (prior_stop). Because baseline adherent and baseline non-adherent status are complementary (base_adh + base_nonadh = 1), only base_nonadh was retained in the longitudinal set and the complementary prior non-use ratio was used in place of base_adh; this preserves 11 longitudinal features while eliminating perfect collinearity between the two baseline-status indicators (Supplementary Table S1). The full variable dictionary and harmonization details are provided in Supplementary Table S1.

### 2.4 Statistical analysis

Two model classes were developed: L2-regularized logistic regression (C=1.0) and random forest (500 trees, minimum leaf samples 20, max features = square root of feature count). All preprocessing (median imputation for continuous variables, mode imputation for categorical variables, standardization) was performed within training folds only; no synthetic oversampling was used, preserving natural event rates (12.6%–22.8%). Within-cohort performance was evaluated by 5-fold × 5-repeat nested cross- validation; hyperparameters were fixed a priori as prespecified defaults (no data-driven tuning), and each outer-fold model was scored only on held-out folds. We report out-of-fold ROC-AUC, Brier score, and calibration intercept and slope (ideal values 0 and 1).

External transport: models fitted on the full CHARLS sample were applied directly to SHARE and HRS without retraining or recalibration, reporting external AUC, Brier score and calibration metrics. To distinguish whether cross-sectional failure reflected China–Europe distance rather than intrinsic non- transportability, we additionally performed leave-one-country-out (LOCO) extrapolation within SHARE.

Simple rule comparators: three deliberately simple rules were evaluated on the same participants and folds: (T0) the baseline carry-forward rule (LOCF; equivalent to a two-state first-order Markov model); (T1) the prior non-use count rule; and (T2) the 3-feature parsimonious medication-history score fit as logistic regression without demographic, clinical or examination information. Non-inferiority of T2 versus the full 32-feature logistic model was tested at a prespecified margin δ=0.03 AUC, using paired- bootstrap confidence intervals (2,000 resamples) on the same out-of-fold predictions.

Country-level meta-analysis: within SHARE, countries with n≥300 and ≥30 outcome events were retained (18 countries). CHARLS-trained models were transported to each country; AUC standard errors were estimated by the Hanley–McNeil method [25], pooled with DerSimonian–Laird random- effects meta-analysis [26], and heterogeneity quantified by I² [12,26]. Inverse-variance weighted univariable meta-regression and Spearman correlations related transported AUC and calibration slope to World Bank indicators (GDP per capita, out-of-pocket expenditure share, treatment coverage, physician density).

Interpretability: SHAP (SHapley Additive exPlanations) was applied to the CHARLS full 32-feature random-forest model, and univariable AUCs were computed for each medication-history feature to identify dominant predictors [27]. Temporal external validation was additionally performed by refitting on earlier baseline waves and predicting the most recent available wave (SHARE 2017→2020, n=2,851; HRS 2018→2020, n=211, exploratory). Analyses used Python (scikit-learn and SHAP). All analysis code is available upon request.

Figure 1 presents the overall study design, including the harmonized cohorts, prospective outcome definition, hierarchical feature sets, model development and validation, simple rule comparators, and the 18-country mechanism analysis.

**Figure 1.**
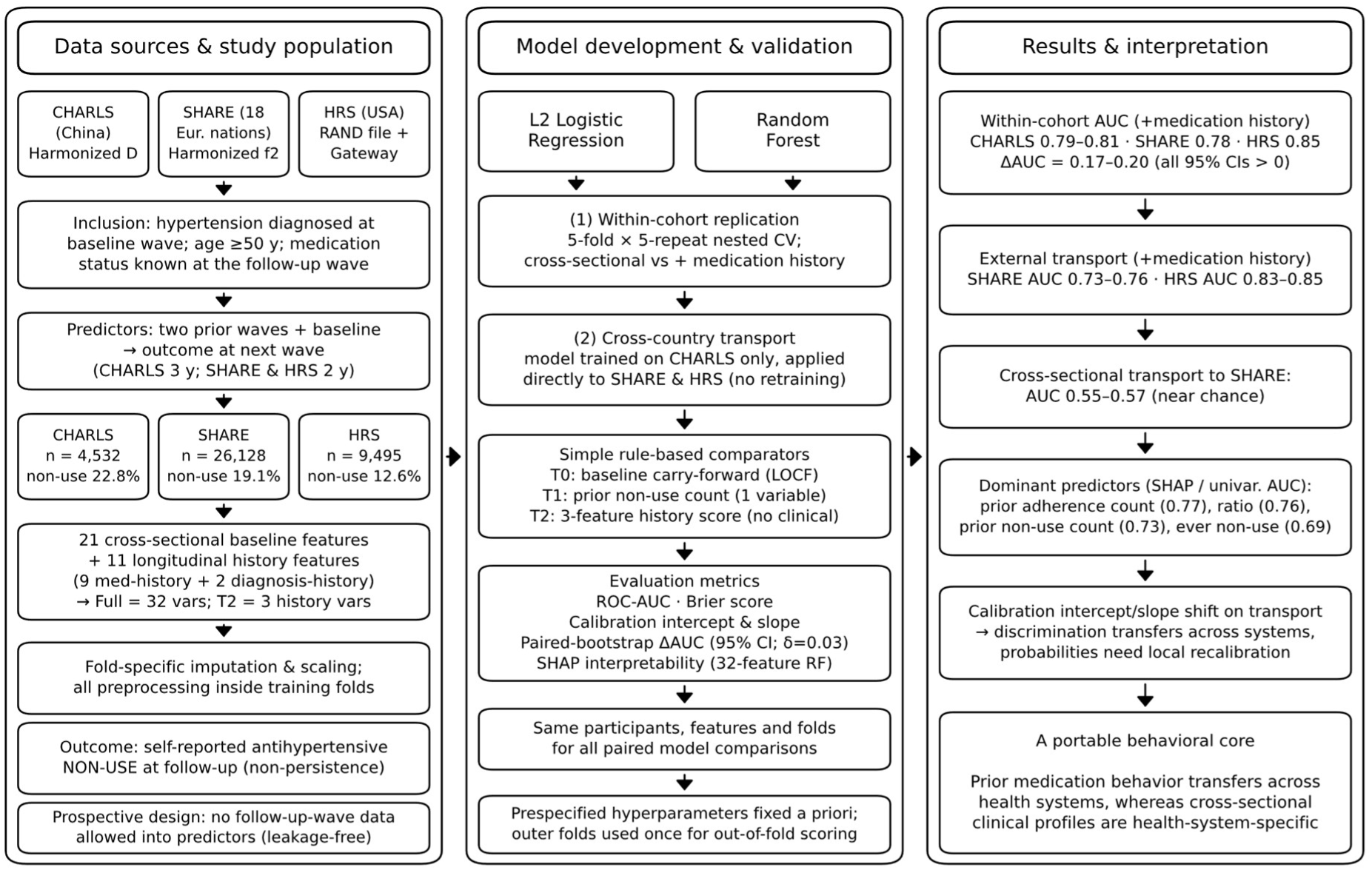
Overall study design: three harmonized prospective cohorts, prospective outcome definition, hierarchical feature sets (cross-sectional 21 / full 32 / T2 parsimonious 3), model development and nested cross- validation, direct cross-country transport, simple rule comparators, and 18-country mechanism analysis.

## 3. Results

### 3.1 Cohort characteristics

Baseline characteristics of the three analytic cohorts are shown in Table 1. A total of 40,155 adults with doctor-diagnosed hypertension were included. CHARLS participants were youngest (mean age 64.3 years, 45.0% male); SHARE participants were oldest (69.6 years, 43.4% male); HRS was intermediate (67.6 years, 41.7% male). Treatment non-persistence was highest in CHARLS (22.8%) and lowest in HRS (12.6%). Baseline antihypertensive use was 80.2% (CHARLS), 84.0% (SHARE) and 87.8% (HRS).

**Table 1.** Baseline characteristics of the three analytic cohorts.

| **Characteristic** | **CHARLS (China)** | **SHARE (Europe)** | **HRS (USA)** |
|---|---|---|---|
| **Participants, n** | 4,532 | 26,128 | 9,495 |
| **Age, years, median (IQR)** | 64 (58–71) | 70 (63–76) | 68 (61–75) |
| **Male, n (%)** | 2,039 (45.0%) | 11,339 (43.4%) | 3,959 (41.7%) |
| **Treatment non-persistence, %** | 22.8 | 19.1 | 12.6 |
| **Baseline antihypertensive use, %** | 80.2 | 84.0 | 87.8 |
| **Diabetes, n (%)** | 1,015 (22.4%) | 5,977 (22.9%) | 2,754 (29.0%) |
| **Heart disease, n (%)** | 822 (18.1%) | 5,686 (21.8%) | 2,849 (30.0%) |
| **Stroke, n (%)** | 363 (8.0%) | 2,012 (7.7%) | 1,289 (13.6%) |
| **Depressive symptoms, n (%)** | 1,849 (40.8%) | 5,487 (21.0%) | 2,279 (24.0%) |
| **Baseline non-use, %** | 19.8 | 16.0 | 12.2 |
| **Prior discontinuation, %** | 17.5 | 14.3 | 9.8 |

### 3.2 Within-cohort replication: medication history dominates

In all three cohorts, adding medication history produced large and statistically significant gains in discrimination, with ΔAUC of 0.17–0.20 and no confidence interval crossing zero (Table 2). The full random-forest model reached AUC 0.809 (CHARLS), 0.781 (SHARE) and 0.850 (HRS), with Brier scores of 0.133, 0.120 and 0.069. The cross-sectional model alone performed only modestly above chance (AUC 0.59–0.67). The parsimonious 3-feature medication-history score (T2) alone reached AUC 0.75–0.83, capturing most of the full model’s discrimination.

**Table 2.** Within-cohort nested cross-validation: cross-sectional vs full (cross-sectional + medication history) vs T2 parsimonious score.

| **Model / feature set** | **CHARLS AUC (Brier)** | **SHARE AUC (Brier)** | **HRS AUC (Brier)** |
|---|---|---|---|
| **Cross-sectional 21 features (LR)** | 0.590 (0.173) | 0.602 (0.151) | 0.665 (0.105) |
| **Cross-sectional 21 features (RF)** | 0.631 (0.169) | 0.609 (0.150) | 0.674 (0.105) |
| **T2 parsimonious 3 features (LR)** | 0.769 (0.143) | 0.748 (0.122) | 0.828 (0.069) |
| **Full 32 features (LR)** | 0.792 (0.137) | 0.776 (0.121) | 0.847 (0.069) |
| **Full 32 features (RF)** | 0.809 (0.133) | 0.781 (0.120) | 0.850 (0.069) |

### 3.3 Cross-country transport: history transfers, cross-sectional does not

The full CHARLS-trained model transported well: AUC 0.73–0.76 in SHARE and 0.83–0.85 in HRS (Table 3). In contrast, the cross-sectional model collapsed to near chance when transported to SHARE (AUC 0.55–0.57) and remained only modest in HRS (AUC 0.62–0.64). Calibration intercepts and slopes deviated from ideal values (0 and 1), particularly for the random forest, indicating that transported discrimination does not imply transported probabilities; local recalibration is required before individual risk thresholds are applied.

**Table 3.** Direct transport of CHARLS-trained models to SHARE and HRS (no retraining)

| **Target cohort** | **Model** | **Cross-sectional AUC** | **Full model AUC** | **Full model Brier** | **Calibration intercept** | **Calibration slope** |
|---|---|---|---|---|---|---|
| SHARE | LR | 0.554 | 0.735 | 0.133 | −0.217 | 0.888 |
| SHARE | RF | 0.566 | 0.753 | 0.128 | 0.271 | 1.184 |
| HRS | LR | 0.616 | 0.833 | 0.083 | −0.177 | 1.175 |
| HRS | RF | 0.640 | 0.847 | 0.079 | 0.653 | 1.710 |

**Figure 2.**
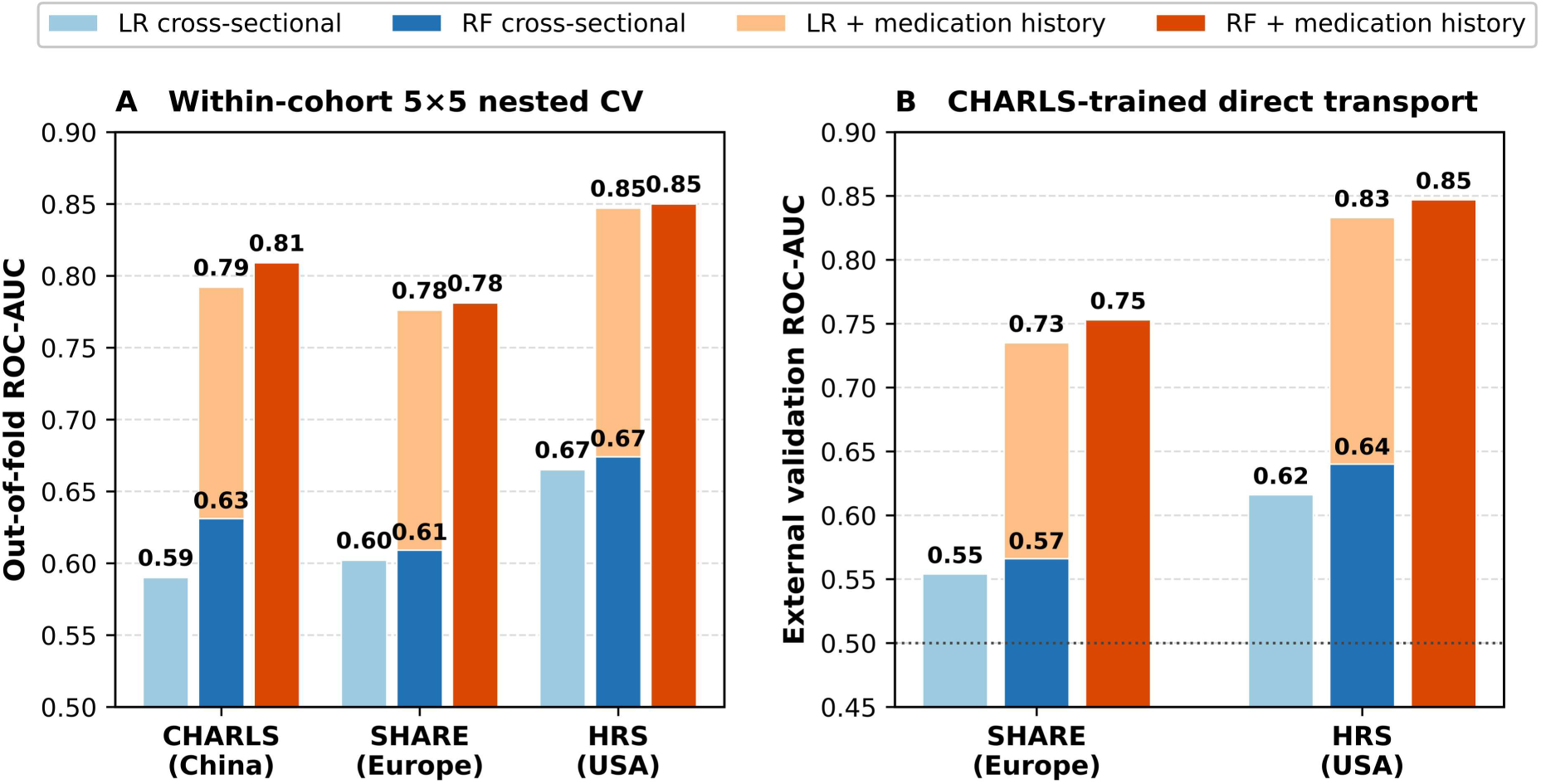
Within-cohort replication (A) and direct cross-country transport (B) of cross-sectional vs full (medication-history) models.

### 3.4 Simple rule comparators and non-inferiority

The single most predictive piece of information was whether the patient was taking medication at baseline: the baseline carry-forward rule (T0) alone achieved AUC 0.70–0.80 (Table 4). The 3-feature parsimonious medication-history score (T2), which contains no demographic, clinical or examination information, reached AUC 0.75–0.83 and captured most of the full model’s discrimination. The full model exceeded T0 by only 0.05–0.08 AUC, and exceeded T2 by 0.018–0.027 AUC (Table 4). At the prespecified margin δ=0.03, T2 was non-inferior to the full 32-feature logistic model in all three within- cohort comparisons (ΔAUC 0.018–0.027; all upper 95% CI bounds <0.03) and in both transport targets, even slightly outperforming the full logistic model when transported from CHARLS to SHARE (ΔAUC −0.011, 95% CI −0.016 to −0.005; Figure 3). The random forest added ∼0.02–0.03 AUC over T2 in within-cohort comparisons.

**Table 4.** Simple rule-based reference comparators vs full models (out-of-fold AUC; non-inferiority margin δ=0.03 for T2 vs full logistic model)

| **Cohort** | **T0 LOCF (baseline status)** | **T1 prior non-use count** | **T2 medication-history score (3 features)** | **Cross-sectional ML (RF)** | **Full ML (LR)** | **Full ML (RF)** |
|---|---|---|---|---|---|---|
| CHARLS | 0.738 | 0.726 | 0.769 | 0.631 | 0.792 | 0.809 |
| SHARE | 0.698 | 0.736 | 0.748 | 0.609 | 0.776 | 0.781 |
| HRS | 0.797 | 0.805 | 0.828 | 0.674 | 0.847 | 0.850 |

**Figure 3.**
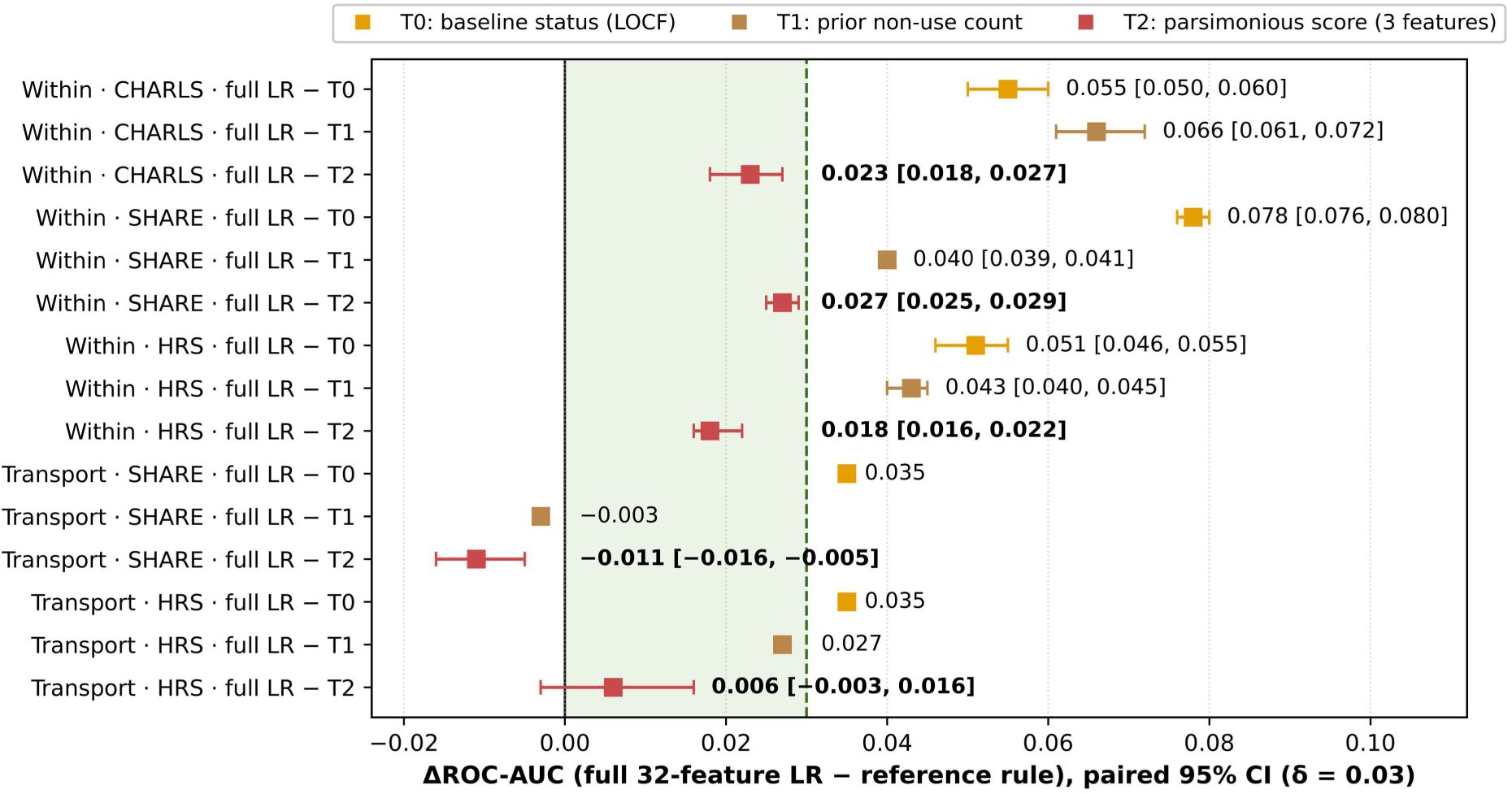
Incremental discrimination of the full model over simple rule-based reference comparators (paired- bootstrap 95% CI; shaded band = non-inferiority margin δ=0.03).

### 3.5 18-country mechanism: where and why history models work

Across 18 European countries, the cross-sectional model was uniformly near chance (pooled AUC 0.55; I²=30%), and even leave-one-country-out transport within SHARE — using the same language, instrument and continent — yielded only 0.59 (I²=23%). This demonstrates that the failure of cross- sectional predictors is not an artefact of China–Europe distance but an intrinsic property of those predictors for this outcome. Medication-history models, by contrast, pooled at 0.74–0.76 (I²=82–87%), with country-level heterogeneity driven substantially by health-system characteristics (Table 5; Figure 4).

**Figure 4.**
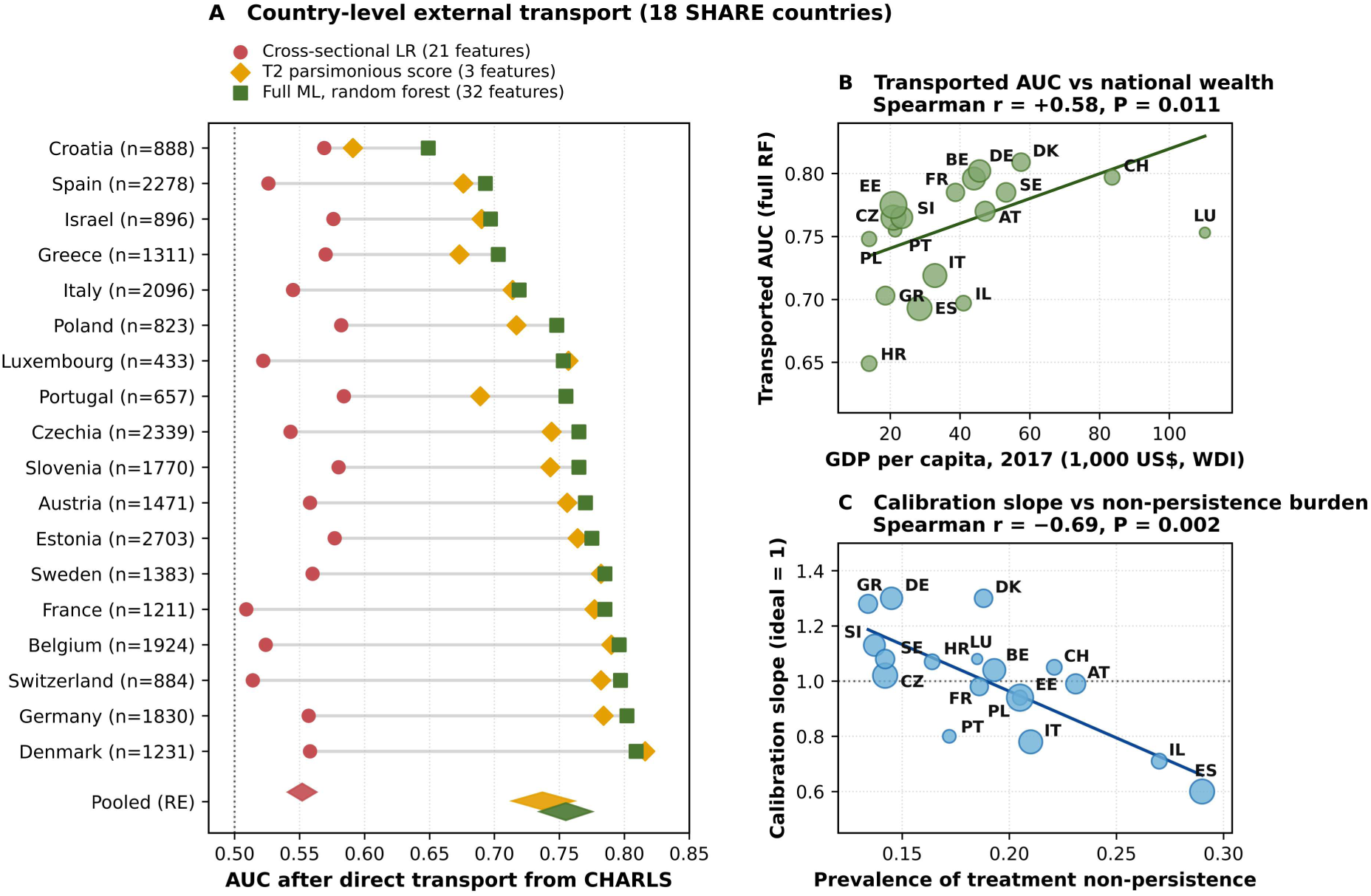
Country-level external transport across 18 SHARE countries (A), and relationships of transported discrimination with national wealth (B) and of calibration slope with non-persistence burden (C).

**Table 5.** Random-effects pooled transport performance across 18 SHARE countries and meta- regression of health-system moderators.

| **Method (CHARLS-trained → direct transport)** | **Pooled AUC** | **95% CI** | **I²** |
|---|---|---|---|
| **Cross-sectional LR (21 features)** | 0.552 | 0.541–0.563 | 30% |
| T0 first-order persistence rule (LOCF) | 0.698 | 0.675–0.721 | 85% |
| T2 parsimonious medication-history score | 0.737 | 0.714–0.761 | 87% |
| **Full model LR (32 features)** | 0.735 | 0.713–0.756 | 84% |
| **Full model random forest** | 0.755 | 0.735–0.775 | 82% |
| Reference: SHARE LOCO, cross-sectional LR | 0.590 | 0.580–0.601 | 23% |
| **Reference: SHARE LOCO, full LR** | 0.762 | 0.739–0.786 | 88% |

Inverse-variance meta-regression showed that transported discrimination was stronger in wealthier countries: full random-forest AUC correlated with log GDP per capita (Spearman r=0.58, P=0.011; weighted R²≈0.22). Calibration showed an even more robust pattern: calibration slope declined with non-persistence burden (r=−0.69, P=0.002) and with out-of-pocket health expenditure share (r=−0.51, P=0.032), and increased with baseline treatment coverage (r=0.62, P=0.006). These findings indicate that history-based models discriminate best in high-resource, high-coverage systems, but that predicted probabilities require local recalibration wherever non-persistence is common (Supplementary Tables S4–S5).

### 3.6 Temporal external validation

Refitting the models on earlier baseline waves and predicting the most recent available wave confirmed the main findings. In SHARE, the full logistic model predicted non-persistence in 2020 (n=2,851; event rate 18.9%) with AUC 0.871 (RF 0.870); the T2 score achieved AUC 0.866 (RF 0.864). In HRS, the full logistic model predicted non-persistence in 2020 (n=211; event rate 28.9%) with AUC 0.908 (RF 0.944); the T2 score achieved AUC 0.910 (RF 0.948). The HRS temporal analysis is exploratory because of the small number of participants with complete harmonized data in the 2020 wave (Supplementary Figure S5).

### 3.7 Dominant predictors

The dominant predictors were medication-history variables: baseline non-use, prior non-use ratio and prior discontinuation contributed the largest shares of model output variance; univariable AUCs were 0.726 (prior non-use count), 0.690 (ever prior non-use) and 0.608 (baseline non-use) in CHARLS (Table 6). SHAP analysis of the CHARLS full 32-feature random-forest model ranked the persistent-wave count, prior non-use ratio and ever-persistent indicator highest by mean |SHAP value|; among cross- sectional features, the largest SHAP contributions were BMI, age and marital status, whereas clinical comorbidity and care-contact indicators (diagnosed heart disease, diabetes, stroke, and recent doctor visits) showed inverse univariable associations with non-persistence (univariable AUC 0.454–0.490) and near-zero SHAP contributions (mean |SHAP| ≤ 0.0015) once medication history was included (Supplementary Table S7).

**Table 6.** Dominant predictors in the full model (CHARLS n=4,532): univariable AUC and SHAP mean |contribution| from the 32-feature random-forest model. *For the protective adherence-count features (n_adh and ever_adh), univariable AUCs are presented as 1 - A UC (0.774 and 0.666) to facilitate magnitude comparison; for all other features, raw univariable AUCs with respect to non-persistence are shown (values <0.50 indicate an inverse/protective association with non-persistence).*

| **Feature** | **Type** | **Univariate AUC** | **Mean \|SHAP\| (RF)** | **Direction** |
|---|---|---|---|---|
| **Prior non-use count (n_nonadh)** | Medication history | 0.726 | 0.023 | More prior non-use → higher risk |
| **Prior non-use ratio (nonadh_ratio)** | Medication history | 0.761 | 0.037 | Higher ratio → higher risk |
| **Ever prior non-use (ever_nonadh)** | Medication history | 0.690 | 0.020 | Any prior non-use → higher risk |
| **Baseline non-use (base_nonadh)** | Medication history | 0.608 | 0.008 | Baseline non-use → higher risk |
| **Prior adherence count (n_adh)** | Medication history | 0.774 | 0.042 | More prior adherence → lower risk |
| **BMI** | Cross-sectional | 0.410 | 0.014 | Higher BMI → lower risk |
| **Age (years)** | Cross-sectional | 0.462 | 0.010 | Older → lower risk |
| **Prior discontinuation (prior_stop)** | Medication history | 0.540 | 0.013 | Prior stop → higher risk |

**Figure 5.**
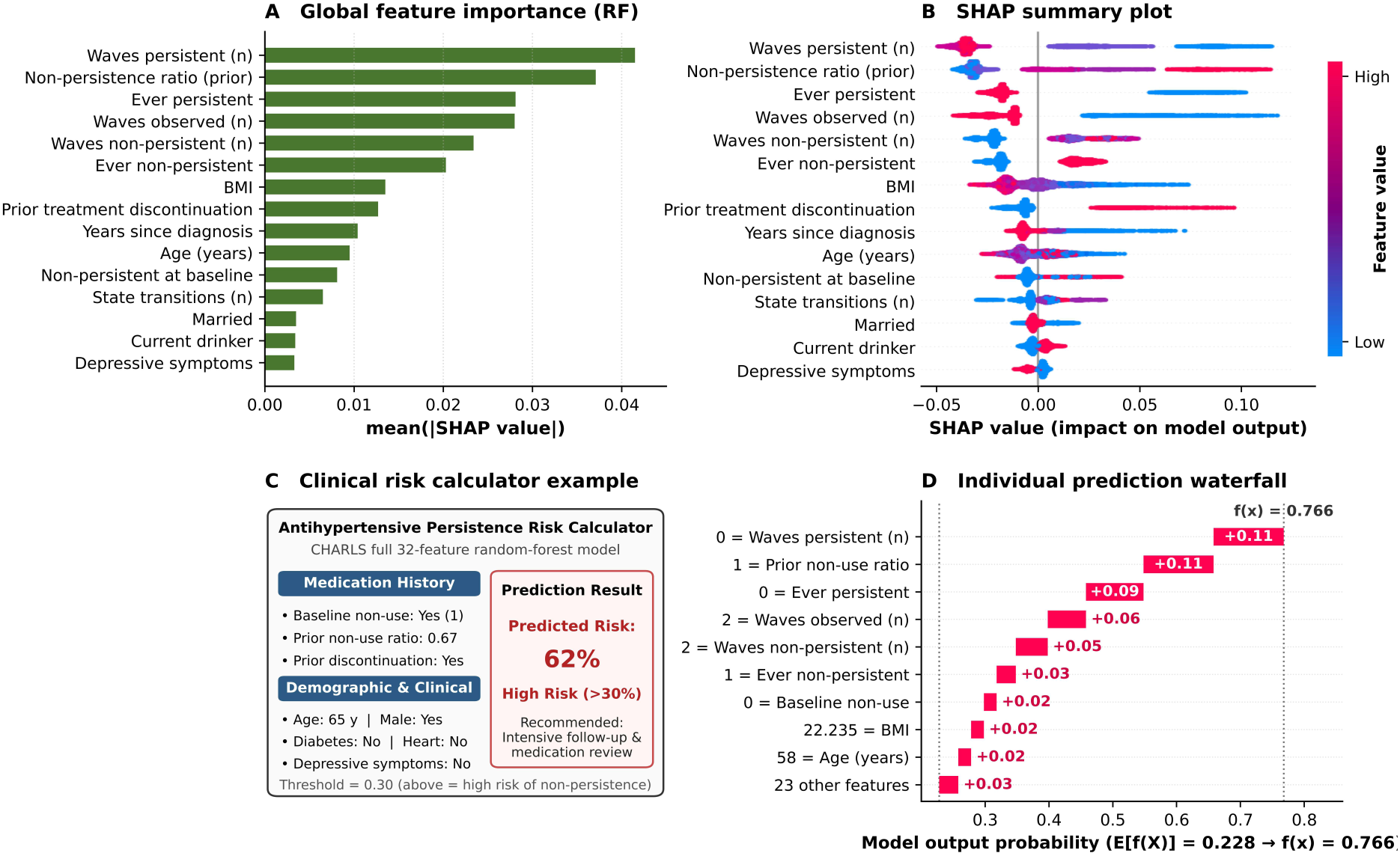
SHAP analysis of the CHARLS full 32-feature random-forest model: (A) mean |SHAP| bar plot; (B) summary bee-swarm plot; (C) prediction calculator example; (D) single-sample waterfall plot.

## 4. Discussion

### 4.1 Main findings

Across three harmonized prospective cohorts spanning China, 18 European countries and the USA, a parsimonious core of medication-history variables predicted future antihypertensive non-persistence far better than cross-sectional clinical profiles, transferred across health systems without retraining, and was non-inferior to a full 32-feature ML model. Cross-sectional clinical profiles performed only modestly above chance within cohorts and collapsed to near chance when transported across systems. Complex ML added little over a one-question baseline carry-forward rule or a 3-variable history score.

### 4.2 Why medication history transfers and clinical profiles do not

Medication history captures an individual’s longitudinal behavioral trajectory — a patient who has previously interrupted or inconsistently taken antihypertensive therapy is substantially more likely to discontinue again — and this behavioral signal is conceptually invariant across healthcare delivery models [15,17,28,29]. Conversely, cross-sectional clinical profiles function largely as proxies for local healthcare utilization and management intensity. In our univariable and SHAP analyses (Supplementary Table S7), diagnosed heart disease, diabetes, prior stroke, and recent physician contact acted in the protective direction (raw univariable AUC 0.454–0.490) and contributed minimally once medication history was controlled, identifying patients who are already integrated into clinical follow-up rather than representing universal biological drivers of persistence. Because diagnostic thresholds, primary- care gatekeeping, and specialist referral pathways vary markedly across China, Europe, and the USA, coefficients learned for these cross-sectional proxies encode system-specific care cascades and consequently collapse upon external transport (pooled AUC 0.552 across 18 European countries). Even under leave-one-country-out validation within Europe using the identical SHARE instrument, cross- sectional models yielded a pooled AUC of only 0.590 (I² = 23%), confirming that their poor transportability is an intrinsic property of cross-sectional clinical features rather than an artefact of geographic distance between China and Europe.

### 4.3 Comparison with previous work

Our results align with and substantially extend three bodies of recent literature. First, in pharmacoepidemiology, longitudinal trajectory studies of antihypertensive and statin therapy have shown that prior refill adherence and early treatment gaps outperform baseline sociodemographic and comorbidity variables in single-country cohorts [15,16,28,29]. While recent single-country ML studies using CHARLS reported cross-sectional AUCs around 0.60–0.67 [16], we demonstrate across three continents that incorporating just two prior waves of medication history elevates AUC to 0.78–0.85. Second, our prespecified non-inferiority findings corroborate systematic methodological evaluations by Dhiman et al. and Collins et al., which show that on structured clinical and epidemiological data, complex ML algorithms rarely provide clinically meaningful discrimination gains over parsimonious regression scores once data leakage is prevented [11,12,29,30]. In fact, during China-to-Europe transport, our 3-feature T2 score outperformed the 32-feature logistic model because the additional 21 cross-sectional variables introduced domain-shift noise. Third, our 18-country meta-regression bridges individual-level prediction transportability with macro-level health-system performance [1,28,31,32]. Consistent with multinational cardiovascular risk validation frameworks such as SCORE2-Diabetes and Riley et al.’s internal–external validation methodology [12,28,31], we found that while ranking discrimination transferred robustly (particularly in wealthier nations with higher log GDP per capita, r=0.58, P=0.011), probability calibration slopes degraded systematically in countries with higher non- persistence burden (r=−0.69, P=0.002), lower treatment coverage (r=0.62, P=0.006), and higher out-of- pocket expenditure shares (r=−0.51, P=0.032) [6,33]. In high cost-sharing systems, structural financial barriers override individual behavioral propensity, compressing or shifting predicted risk distributions and necessitating local recalibration of baseline hazard and slope before applying absolute probability thresholds [28,31].

### 4.4 Implications for clinical practice and health policy

The practical implication is that risk stratification for antihypertensive non-persistence can be based on two or three routinely available questions — “Are you currently taking your medication?”, “How often have you missed it in the past?”, “Have you ever stopped it?” — without requiring complex ML, cross- sectional biomarkers or country-specific recalibration for ranking purposes. In resource-rich, high- coverage systems, history-based models achieve strong discrimination (AUC 0.83–0.85 in HRS). Where non-persistence is common and out-of-pocket costs are high, predicted probabilities are systematically miscalibrated and require local recalibration; the ranking, however, remains informative. This supports the design of lightweight screening tools in primary care and of adherence-support programs targeted to individuals identified by history-based risk scores [7,8].

### 4.5 Strengths and limitations

Strengths include three harmonized cohorts across continents, a prospective outcome definition, nested cross-validation, direct external transport, prespecified non-inferiority testing against simple rules, and country-level meta-analysis. Limitations should be acknowledged. First, medication use was self- reported; misclassification is possible, although self-reported non-use is a conservative endpoint with clinical meaning. Second, follow-up intervals differed across cohorts (2 vs 3 years), which may affect persistence levels; our within-cohort comparisons are unaffected, but cross-cohort absolute comparisons should be interpreted cautiously. Third, the primary outcome was defined among all diagnosed participants regardless of baseline medication status; in a sensitivity analysis restricted to participants actively using antihypertensives at baseline (71.5%–87.5% of each cohort; Supplementary Table S8), incident discontinuation rates were lower (5.1%–11.8%) and overall AUCs were correspondingly more modest (full-model AUC 0.642–0.674), as expected for a harder prediction problem. Because restricting to baseline users renders the baseline non-use indicator constant (base_nonadh = 0 for all individuals), the 3-feature T2 score effectively reduces to a 2-variable score (AUC 0.590–0.634), widening its gap against the full 32-feature model (ΔAUC 0.023 in CHARLS, 0.058 in SHARE, and 0.051 in HRS); nevertheless, longitudinal medication-history models still consistently outperformed cross-sectional clinical profiles. Fourth, harmonization across cohorts limited the cross-sectional feature set to 21 variables, excluding candidate predictors such as measured blood pressure, medication class, or out-of-pocket cost, which may matter within single systems. Fifth, the outcome does not distinguish intermittent non-adherence from complete discontinuation. Finally, transportability was assessed from one training system (China) to European and US systems; transport in the reverse direction was not tested.

## 5. Conclusions

In three harmonized prospective cohorts, a 3-feature medication-history score predicted future antihypertensive non-persistence with AUC 0.75–0.83, transferred across health systems without retraining, and was non-inferior to a full 32-feature machine-learning model. Cross-sectional clinical profiles predicted only modestly within cohorts and failed across systems. Health-system context — national wealth, treatment coverage and out-of-pocket burden — modulates transported discrimination and calibration, indicating that history-based risk scores can be deployed broadly for ranking, but require local recalibration when used for individual probability thresholds.

## Data Availability

The data supporting the findings of this study are available from the respective cohort repositories to registered researchers. CHARLS data can be accessed at https://charls.pku.edu.cn/; SHARE data can be accessed at https://share-eric.eu/data/data-access; and HRS data can be accessed at https://hrsdata.isr.umich.edu/. Harmonized files are available through the Gateway to Global Aging Data (https://g2aging.org). World Bank indicators are publicly accessible at https://data.worldbank.org. Analysis code is available from the corresponding author upon reasonable request.

## List of abbreviations

ADL: activities of daily living;
AUC: area under the receiver operating characteristic curve
BMI: body mass index
CHARLS: China Health and Retirement Longitudinal Study
CI: confidence interval
CV: cross-validation
DCA: decision curve analysis
GDP: gross domestic product
HRS: Health and Retirement Study
IADL: instrumental activities of daily living
IQR: interquartile range
LOCF: last observation carried forward
LOCO: leave-one-country-out
LR: logistic regression
ML: machine learning
OOF: out-of-fold
OOP: out-of-pocket
PR: precision-recall
RF: random forest
SHAP: SHapley Additive exPlanations
SHARE: Survey of Health, Ageing and Retirement in Europe
WHO: World Health Organization

## Declarations

### Ethics approval and consent to participate

The present study was a secondary analysis of de-identified data and involved no direct contact with participants; no additional ethics approval or registration was required at the authors’ institution. The CHARLS protocol was approved by the Institutional Review Board at Peking University (IRB00001052-11015). The HRS protocol was approved by the University of Michigan Health Sciences/Behavioral Sciences Institutional Review Board. The SHARE protocol was approved by the Ethics Committee of the University of Mannheim/the Max Planck Society. Participants provided informed consent in accordance with the approved study procedures.

### Consent for publication

Not applicable.

### Availability of data and materials

CHARLS, HRS and SHARE data are available to registered researchers subject to the access and terms- of-use requirements of the respective cohort repositories: https://charls.pku.edu.cn/; https://hrsdata.isr.umich.edu/; and https://share-eric.eu/data/data-access. World Bank indicators are publicly available (https://data.worldbank.org). Analysis code is available from the corresponding author on reasonable request.

### Authors’ contributions

DL and SW curated the data, conducted the statistical analyses and prepared the initial draft of the manuscript. TT contributed to the interpretation of the findings and critically revised the manuscript. SW contributed to the critical revision of the manuscript and overall supervision. All authors reviewed and approved the final manuscript. SW is the guarantor and accepts full responsibility for the finished work and the conduct of the study, had access to the data and controlled the decision to publish.

## Acknowledgements

We thank the participants, interviewers, investigators and data-management teams of the CHARLS, the HRS, and the SHARE for making these cohort resources available for scientific research. The CHARLS, HRS and SHARE harmonised data files were made available through the Gateway to Global Aging Data.

## SUPPLEMENTARY MATERIAL

Supplementary Methods, Tables S1–S8, and Figures S1–S5

## S1. Supplementary methods

### S1.1 Data harmonization

All three cohorts were accessed through the Gateway to Global Aging Data harmonized files. Hypertension was defined as self-reported doctor diagnosis. Antihypertensive use was self-reported current use of any antihypertensive medication. The harmonized medication-use variables were rxhibp (CHARLS: rxhibp_c; SHARE: rxhibp; HRS Gateway: r{wave}rxhibp). For HRS, the RAND 2020 longitudinal file was merged with the Gateway medication-use file by hhidpn. Medication status was available for waves 2011–2018 (CHARLS; baseline waves up to 2015 with 3-year follow-up to 2018), 2011–2017 (SHARE) and 2012–2018 (HRS); the temporal validation additionally used SHARE 2020 (n=2,851) and HRS 2020 (n=211). For participants entering at the first available wave of a cohort (with no preceding observation waves), the number of observed waves (n_obs) was set to 1, prior adherent and non-adherent wave counts reflected the baseline status, and state-transition count (n_state_chg) and prior discontinuation (prior_stop) were coded as 0.

### S1.2 Missing values and preprocessing

Continuous variables were imputed with the median and categorical variables with the mode; all imputation was performed within training folds only. Continuous variables were standardized (z-score) within training folds. No synthetic oversampling (e.g., SMOTE) was used, preserving natural event rates (12.6%–22.8%). Binary and ordinal categorical variables were entered directly; tree models did not receive one-hot encoding, and logistic regression used original ordinal coding.

### S1.3 Model specifications

L2-regularized logistic regression used C=1.0 with default solver. Random forest used 500 trees, minimum leaf samples 20, and max features = square root of the feature count, without a maximum depth. Hyperparameters were fixed a priori as prespecified defaults; no data-driven tuning was performed, and each outer-fold model was scored only on held-out folds. All random seeds were fixed to 12345 for reproducibility.

### S1.4 Non-inferiority testing

A margin of δ=0.03 AUC was prespecified as the minimum clinically meaningful difference in discrimination for chronic-disease risk prediction models. The parsimonious 3-feature medication- history score (T2) was declared non-inferior to the full 32-feature logistic model when the upper bound of the 95% CI of (full-model AUC − T2 AUC) fell below 0.03. Confidence intervals were obtained by paired bootstrap resampling (2,000 resamples) on the same out-of-fold predictions.

### S1.5 Meta-analysis

Country-level AUC standard errors were estimated by the Hanley–McNeil method. Pooled estimates used DerSimonian–Laird random-effects meta-analysis; heterogeneity was quantified by I². Meta- regression used inverse-variance weighted univariable linear regression of country-level AUC and calibration slope on each moderator. World Bank indicators (GDP per capita, out-of-pocket expenditure as % of current health expenditure, physicians per 1,000 people) were extracted for 2017 (or the most recent available year) via the World Bank API.

## S2. Variable dictionary and harmonization notes

Table S1 lists all 32 features used in the full model: 21 cross-sectional baseline features and 11 longitudinal medication-history features. Because baseline adherent status (base_adh) and baseline non- adherent status (base_nonadh) are complementary indicators (base_adh + base_nonadh = 1), only base_nonadh was retained, and the prior non-use ratio (nonadh_ratio = number of non-adherent waves / number of observed waves) was used in place of base_adh. This preserves 11 longitudinal features while eliminating the perfect collinearity between the two baseline-status indicators.

**Table S1.**
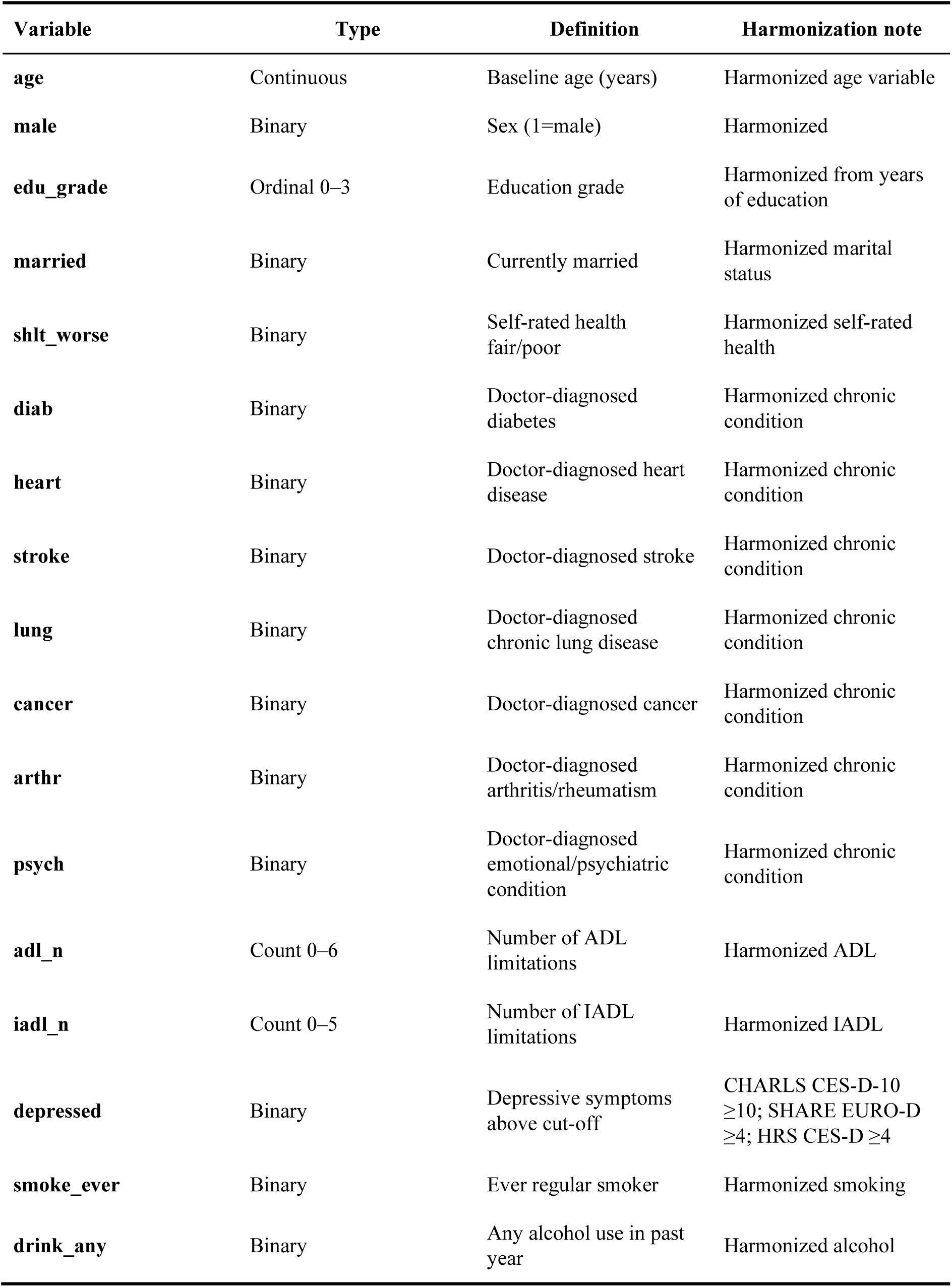
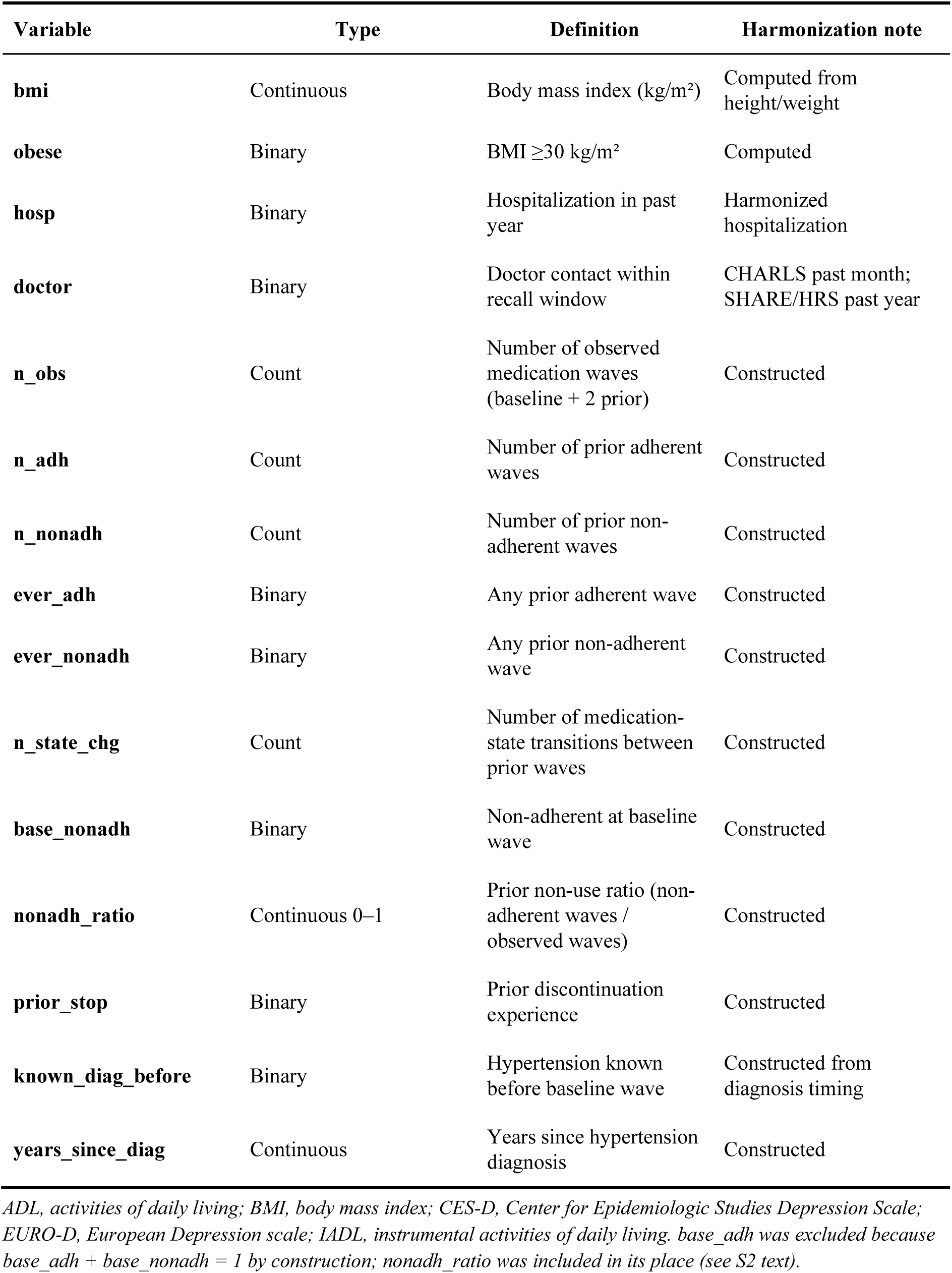
Full variable dictionary: 21 cross-sectional baseline features and 11 longitudinal medication-history features.

| Variable | Type | Definition | Harmonization note |
|---|---|---|---|
| **age** | Continuous | Baseline age (years) | Harmonized age variable |
| **male** | Binary | Sex (1=male) | Harmonized |
| **edu_grade** | Ordinal 0–3 | Education grade | Harmonized from years of education |
| **married** | Binary | Currently married | Harmonized marital status |
| **shlt_worse** | Binary | Self-rated health fair/poor | Harmonized self-rated health |
| **diab** | Binary | Doctor-diagnosed diabetes | Harmonized chronic condition |
| **heart** | Binary | Doctor-diagnosed heart disease | Harmonized chronic condition |
| **stroke** | Binary | Doctor-diagnosed stroke | Harmonized chronic condition |
| **lung** | Binary | Doctor-diagnosed chronic lung disease | Harmonized chronic condition |
| **cancer** | Binary | Doctor-diagnosed cancer | Harmonized chronic condition |
| **arthr** | Binary | Doctor-diagnosed arthritis/rheumatism | Harmonized chronic condition |
| **psych** | Binary | Doctor-diagnosed emotional/psychiatric condition | Harmonized chronic condition |
| **adl_n** | Count 0–6 | Number of ADL limitations | Harmonized ADL |
| **iadl_n** | Count 0–5 | Number of IADL limitations | Harmonized IADL |
| **depressed** | Binary | Depressive symptoms above cut-off | CHARLS CES-D-10 ≥10; SHARE EURO-D ≥4; HRS CES-D ≥4 |
| **smoke_ever** | Binary | Ever regular smoker | Harmonized smoking |
| **drink_any** | Binary | Any alcohol use in past year | Harmonized alcohol |
| **Variable** | **Type** | **Definition** | **Harmonization note** |
| **bmi** | Continuous | Body mass index (kg/m<sup>2</sup>) | Computed from height/weight |
| **obese** | Binary | BMI ≥30 kg/m<sup>2</sup> | Computed |
| **hosp** | Binary | Hospitalization in past year | Harmonized hospitalization |
| **doctor** | Binary | Doctor contact within recall window | CHARLS past month; SHARE/HRS past year |
| **n_obs** | Count | Number of observed medication waves (baseline + 2 prior) | Constructed |
| **n_adh** | Count | Number of prior adherent waves | Constructed |
| **n_nonadh** | Count | Number of prior non-adherent waves | Constructed |
| **ever_adh** | Binary | Any prior adherent wave | Constructed |
| **ever_nonadh** | Binary | Any prior non-adherent wave | Constructed |
| **n_state_chg** | Count | Number of medication-state transitions between prior waves | Constructed |
| **base_nonadh** | Binary | Non-adherent at baseline wave | Constructed |
| **nonadh_ratio** | Continuous 0–1 | Prior non-use ratio (non-adherent waves / observed waves) | Constructed |
| **prior_stop** | Binary | Prior discontinuation experience | Constructed |
| **known_diag_before** | Binary | Hypertension known before baseline wave | Constructed from diagnosis timing |
| **years_since_diag** | Continuous | Years since hypertension diagnosis | Constructed |

*ADL, activities of daily living; BMI, body mass index; CES-D, Center for Epidemiologic Studies Depression Scale; EURO-D, European Depression scale; IADL, instrumental activities of daily living. base\_adh was excluded because base\_adh + base\_nonadh = 1 by construction; nonadh\_ratio was included in its place (see S2 text).*

## S3. Supplementary results: country-level external validation

**Table S2.** Country-level external transport results (CHARLS-trained models → 18 SHARE countries)

| **Country** | **n** | **Events** | **Non-persistence %** | **Cross-sectional LR** | **T0 LOCF** | **T2 score** | **Full LR** | **Full RF** | **Calibration slope** |
|---|---|---|---|---|---|---|---|---|---|
| Austria | 1,471 | 340 | 23.1% | 0.558 | 0.700 | 0.756 | 0.759 | 0.770 | 0.99 |
| Belgium | 1,924 | 371 | 19.3% | 0.524 | 0.753 | 0.790 | 0.773 | 0.796 | 1.04 |
| Switzerland | 884 | 195 | 22.1% | 0.514 | 0.736 | 0.782 | 0.782 | 0.797 | 1.05 |
| Czechia | 2,339 | 332 | 14.2% | 0.543 | 0.693 | 0.744 | 0.755 | 0.765 | 1.02 |
| Germany | 1,830 | 266 | 14.5% | 0.557 | 0.736 | 0.784 | 0.788 | 0.802 | 1.30 |
| Denmark | 1,231 | 231 | 18.8% | 0.558 | 0.776 | 0.816 | 0.801 | 0.809 | 1.30 |
| Estonia | 2,703 | 554 | 20.5% | 0.577 | 0.717 | 0.764 | 0.755 | 0.775 | 0.94 |
| Spain | 2,278 | 660 | 29.0% | 0.526 | 0.615 | 0.676 | 0.675 | 0.693 | 0.60 |
| France | 1,211 | 225 | 18.6% | 0.509 | 0.728 | 0.777 | 0.757 | 0.785 | 0.98 |
| Greece | 1,311 | 176 | 13.4% | 0.570 | 0.673 | 0.673 | 0.673 | 0.703 | 1.28 |
| Croatia | 888 | 146 | 16.4% | 0.569 | 0.591 | 0.591 | 0.622 | 0.649 | 1.07 |
| Israel | 896 | 242 | 27.0% | 0.576 | 0.649 | 0.690 | 0.678 | 0.697 | 0.71 |
| Italy | 2,096 | 441 | 21.0% | 0.545 | 0.655 | 0.714 | 0.706 | 0.719 | 0.78 |
| Luxembourg | 433 | 80 | 18.5% | 0.522 | 0.706 | 0.757 | 0.762 | 0.753 | 1.08 |
| Poland | 823 | 169 | 20.5% | 0.582 | 0.696 | 0.717 | 0.702 | 0.748 | 0.94 |
| Portugal | 657 | 113 | 17.2% | 0.584 | 0.708 | 0.689 | 0.685 | 0.755 | 0.80 |
| Sweden | 1,383 | 196 | 14.2% | 0.560 | 0.732 | 0.782 | 0.772 | 0.785 | 1.08 |
| Slovenia | 1,770 | 242 | 13.7% | 0.580 | 0.699 | 0.743 | 0.750 | 0.765 | 1.13 |

*AUC, area under the ROC curve; LOCF, last observation carried forward; LR, logistic regression; RF, random forest; T2, 3-feature parsimonious medication-history score.*

**Table S3.**
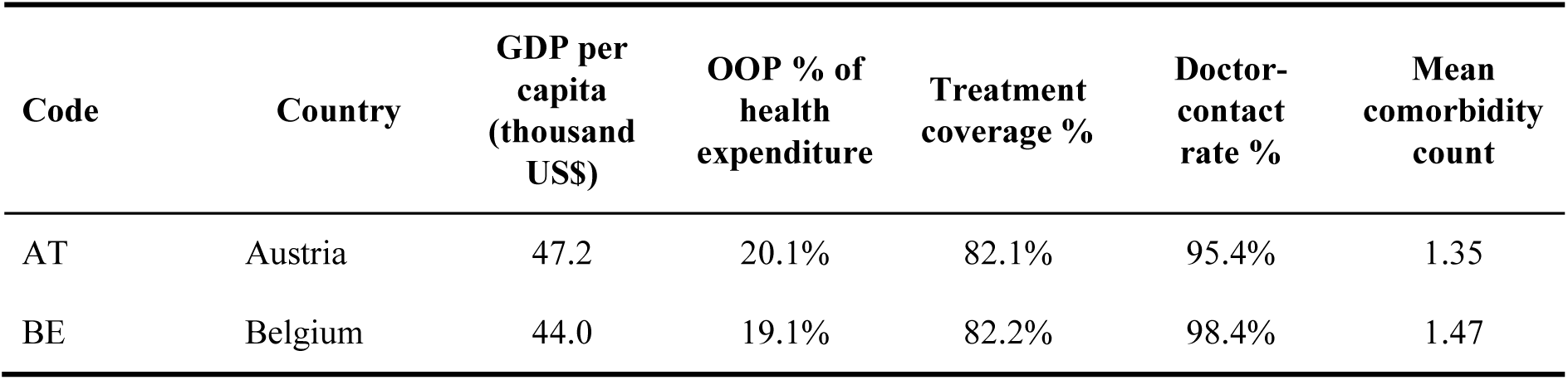
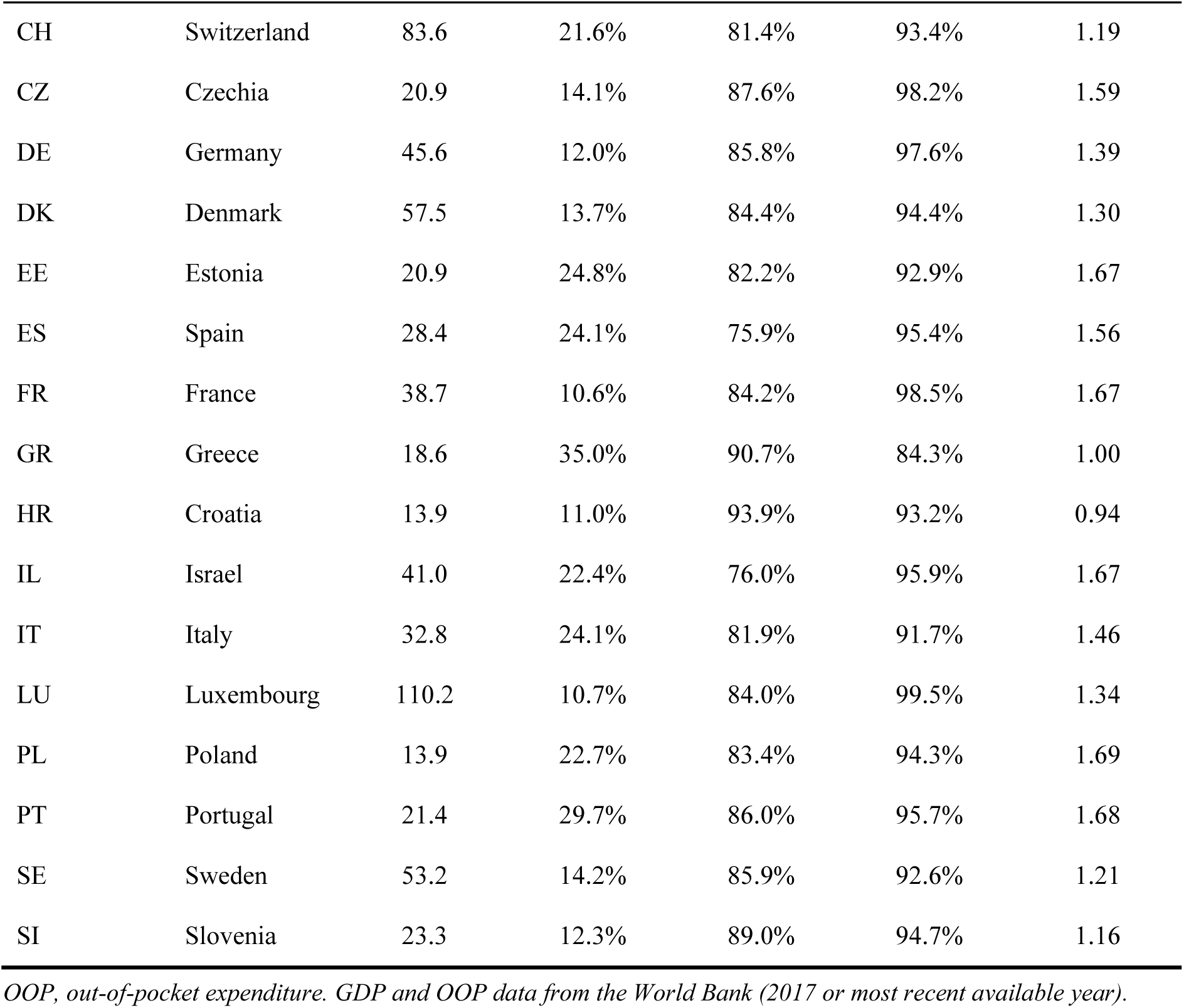
Country-level health-system indicators and meta-regression moderators.

**Table S4.** Full univariable meta-regression results (outcome: transported full-model random-forest AUC)

| **Moderator** | **Slope** | **SE** | **P value** | **Weighted R<sup>2</sup>** | **Spearman r** | **Spearman P** |
|---|---|---|---|---|---|---|
| log(GDP per capita) | +0.041 | 0.020 | 0.054 | 0.22 | +0.58 | 0.011 |
| Non-persistence rate | −0.413 | 0.203 | 0.052 | 0.21 | −0.11 | 0.661 |
| OOP % of health expenditure | −0.003 | 0.002 | 0.078 | 0.15 | −0.33 | 0.185 |
| Baseline treatment coverage | +0.002 | 0.002 | 0.353 | 0.06 | +0.21 | 0.402 |
| Doctor-contact rate | +0.001 | 0.001 | 0.157 | 0.03 | +0.15 | 0.551 |
| Mean comorbidity count | +0.012 | 0.015 | 0.940 | 0.04 | +0.18 | 0.473 |
| Physicians per 1,000 | +0.002 | 0.004 | 0.556 | 0.02 | +0.06 | 0.812 |

*GDP, gross domestic product; OOP, out-of-pocket; SE, standard error.*

**Table S5.** Meta-regression of calibration slope on health-system moderators.

| **Moderator** | **Spearman r** | **P value** | **Interpretation** |
|---|---|---|---|
| **Non-persistence rate** | −0.69 | 0.002 | Higher burden → worse calibration (slope far below 1) |
| **Baseline treatment coverage** | +0.62 | 0.006 | Higher coverage → better calibration (slope toward 1) |
| **OOP % of health expenditure** | −0.51 | 0.032 | Higher cost-sharing → worse calibration |
| **log(GDP per capita)** | +0.29 | 0.239 | Wealthier → trend toward better calibration |
| **Doctor-contact rate** | −0.06 | 0.807 | No significant association |

## S4. Sensitivity analyses: alternative samples and specifications

**Table S6.** Sensitivity analyses: alternative samples and specifications.

| Analysis | Sample | Model | AUC | Description |
|---|---|---|---|---|
| **Primary analysis (harmonized ≥50)** | CHARLS n=4,532 | RF (full) | 0.809 | Three-cohort common feature set |
| Prospective ≥45 (developmental cohort) | CHARLS n=4,295 | RF (full) | 0.772 | Age ≥45, 2015→2018 |
| Prospective ≥45 (developmental cohort) | CHARLS n=4,295 | LR (full) | 0.756 | Age ≥45 |
| Cross-sectional 2018 (developmental cohort) | CHARLS n=7,151 | Best of 8 models | 0.666 | Single wave, LASSO-selected 18 features |
| Cross-sectional 2018, complete cases | CHARLS n=4,701 | LR | 0.670 | No imputation |
| New dimensions (D1 exam + D2 management) | CHARLS n=4,295 | RF | 0.778 | Adding measured blood pressure + community management |
| Univariable: baseline medication status | CHARLS n=4,295 | LOCF rule | 0.692 | 2015 status → 2018 outcome |
| **Univariable: prior non-use count** | CHARLS n=4,532 | Count rule | 0.726 | Univariable AUC |
| SHARE leave-one-country-out (full model) | 18 countries | LR | 0.762 | Pooled, I<sup>2</sup>=88% |
| SHARE leave-one-country-out (cross-sectional) | 18 countries | LR | 0.590 | Pooled, I<sup>2</sup>=23% |

*All sensitivity analyses were consistent with the primary findings: medication-history features dominated, cross-sectional features had limited predictive value, and the baseline carry-forward rule (T0) captured most of the signal. The CHARLS developmental cohort (age ≥45, n=4,295) reproduced the results of the primary harmonized ≥50 three-cohort sample.*

## S5. Complete SHAP feature importance

**Table S7.** Complete SHAP mean |contribution| for the CHARLS full 32-feature model (n=4,532; random forest and logistic regression)

| **Feature** | **Univariate AUC** | **RF mean \|SHAP\|** | **LR mean \|SHAP\|** |
|---|---|---|---|
| **Waves persistent (n_adh)** | 0.774 | 0.0415 | 0.3014 |
| Non-persistence ratio (nonadh_ratio) | 0.761 | 0.0371 | 0.1985 |
| **Ever persistent (ever_adh)** | 0.666 | 0.0281 | 0.2129 |
| **Waves observed (n_obs)** | 0.623 | 0.0280 | 0.2321 |
| **Waves non-persistent (n_nonadh)** | 0.726 | 0.0234 | 0.1782 |
| **Ever non-persistent (ever_nonadh)** | 0.690 | 0.0203 | 0.2560 |
| **BMI** | 0.410 | 0.0135 | 0.0004 |
| **Prior discontinuation (prior_stop)** | 0.540 | 0.0127 | 0.3037 |
| **Years since diagnosis** | 0.380 | 0.0104 | 0.0205 |
| **Age (years)** | 0.462 | 0.0095 | 0.0335 |
| **Baseline non-use (base_nonadh)** | 0.608 | 0.0081 | 0.2180 |
| **State transitions (n_state_chg)** | 0.527 | 0.0065 | 0.1920 |
| **Married** | 0.493 | 0.0035 | 0.0339 |
| **Current drinker** | 0.533 | 0.0034 | 0.0673 |
| **Depressive symptoms** | 0.483 | 0.0033 | 0.0892 |
| **Worsened self-rated health** | 0.469 | 0.0032 | 0.0277 |
| **ADL limitations (n)** | 0.491 | 0.0023 | 0.0552 |
| **Arthritis** | 0.499 | 0.0020 | 0.0578 |
| **Ever smoker** | 0.512 | 0.0016 | 0.0555 |
| **Heart disease** | 0.454 | 0.0015 | 0.0114 |
| **Diabetes** | 0.470 | 0.0014 | 0.0393 |
| **Male** | 0.527 | 0.0014 | 0.0552 |
| **Doctor visit (recall window)** | 0.481 | 0.0013 | 0.0214 |
| **Hospitalized (1y)** | 0.479 | 0.0009 | 0.0130 |
| **Chronic lung disease** | 0.491 | 0.0007 | 0.0229 |
| **Stroke** | 0.490 | 0.0005 | 0.0118 |
| **Psychiatric disease** | 0.507 | 0.0005 | 0.0572 |
| **Education level** | 0.497 | 0.0004 | 0.0232 |
| **Obese (BMI≥30)** | 0.483 | 0.0004 | 0.0392 |
| **IADL limitations (n)** | 0.500 | 0.0004 | 0.0064 |
| **Cancer** | 0.502 | 0.0000 | 0.0202 |
| **Diagnosed before baseline** | 0.500 | 0.0000 | 0.0000 |

*For the protective adherence-count features (n\_adh and ever\_adh), univariable AUCs are presented as 1 − AUC (0.774 and 0.666) to facilitate magnitude comparison; for all other features, raw univariable AUCs with respect to non-persistence are shown (values <0.50 indicate an inverse/protective association with non-persistence). LR mean |SHAP| values are larger than RF values because linear-model SHAP values are not normalized by tree path averaging; rankings within each model are the relevant comparison. The dominant predictors are medication-history variables in both models; cross-sectional features make smaller contributions.*

## S6. Sensitivity analysis restricted to baseline antihypertensive users

**Table S8.** Sensitivity analysis restricted to participants using antihypertensives at baseline (base_adh = 1)

| Cohort | n (baseline users) | % of full sample | Event rate | Model | T2 AUC (Brier) | Full AUC (Brier) | Δ AUC (full − T2) [95% CI] |
|---|---|---|---|---|---|---|---|
| CHARLS | 3,239 | 71.5% | 10.4% | LR | 0.634 (0.090) | 0.659 (0.091) | 0.023 [0.014, 0.033] |
| CHARLS | 3,239 | 71.5% | 10.4% | RF | 0.628 (0.091) | 0.674 (0.090) | 0.044 [0.035, 0.052] |
| SHARE | 21,914 | 83.9% | 11.8% | LR | 0.597 (0.101) | 0.656 (0.099) | 0.058 [0.054, 0.063] |
| SHARE | 21,914 | 83.9% | 11.8% | RF | 0.597 (0.101) | 0.661 (0.099) | 0.064 [0.059, 0.068] |
| HRS | 8,310 | 87.5% | 5.1% | LR | 0.590 (0.048) | 0.642 (0.048) | 0.051 [0.041, 0.062] |
| HRS | 8,310 | 87.5% | 5.1% | RF | 0.584 (0.048) | 0.648 (0.048) | 0.063 [0.053, 0.073] |

*When the analysis was restricted to participants already using antihypertensive medication at baseline (base\_adh = 1), incident discontinuation rates fell to 5.1%–11.8% and overall AUCs were lower than in the full sample. Because base\_nonadh is invariant (0 for all baseline users) in this subgroup, T2 reduces to two active history variables, resulting in a larger discrimination gain from the full 32-feature model (ΔAUC 0.023–0.058 for LR) while still confirming the predictive value of prior longitudinal history. Percentages of baseline users are expressed relative to the full analytic sample; the corresponding baseline-use proportions in Table 1 (80.2% for CHARLS, 84.0% for SHARE, 87.8% for HRS) were computed among participants with known baseline medication status (in CHARLS, 3,239/4,038 = 80.2%, because 494 participants lacked baseline medication-use data and were excluded from that denominator).*

## S7. Supplementary figures

**Figure S1.**
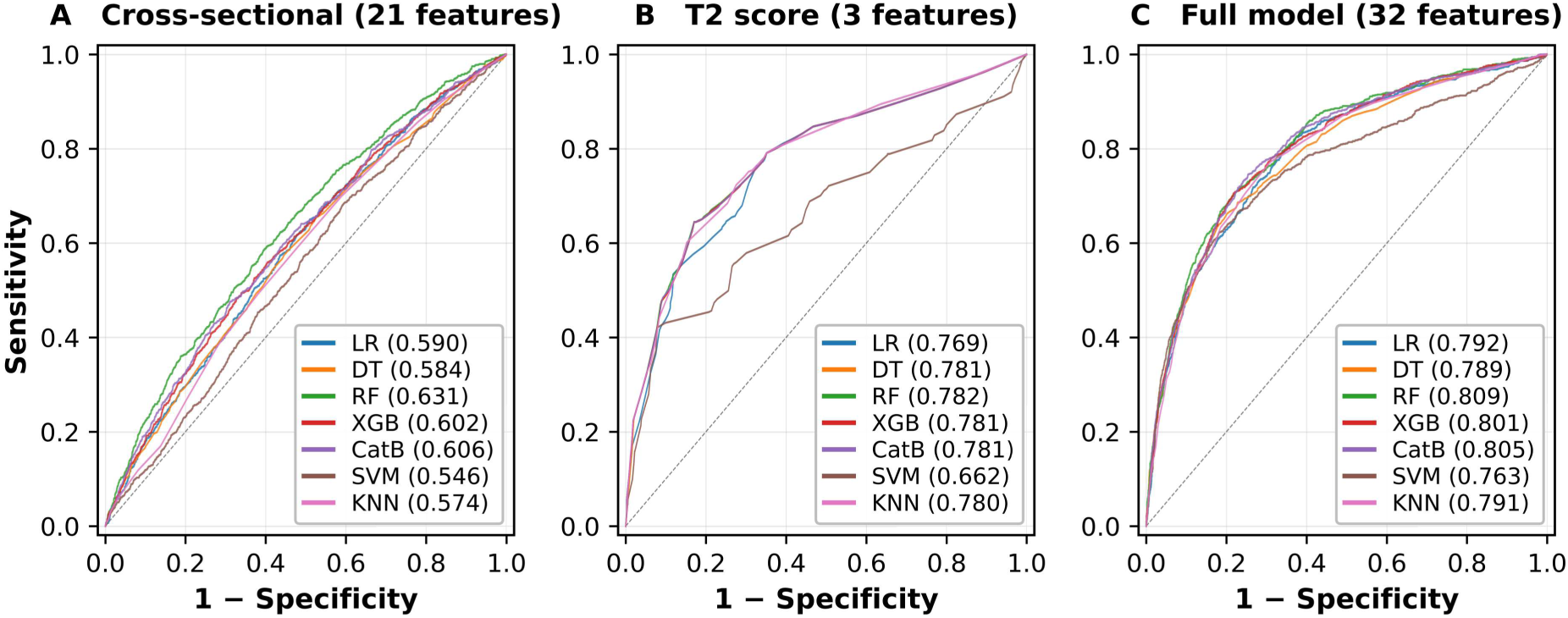
ROC curves for the three feature sets (cross-sectional 21 / T2 parsimonious 3 / full 32) across seven model classes (CHARLS, nested 5×5 out-of-fold).

**Figure S2.**
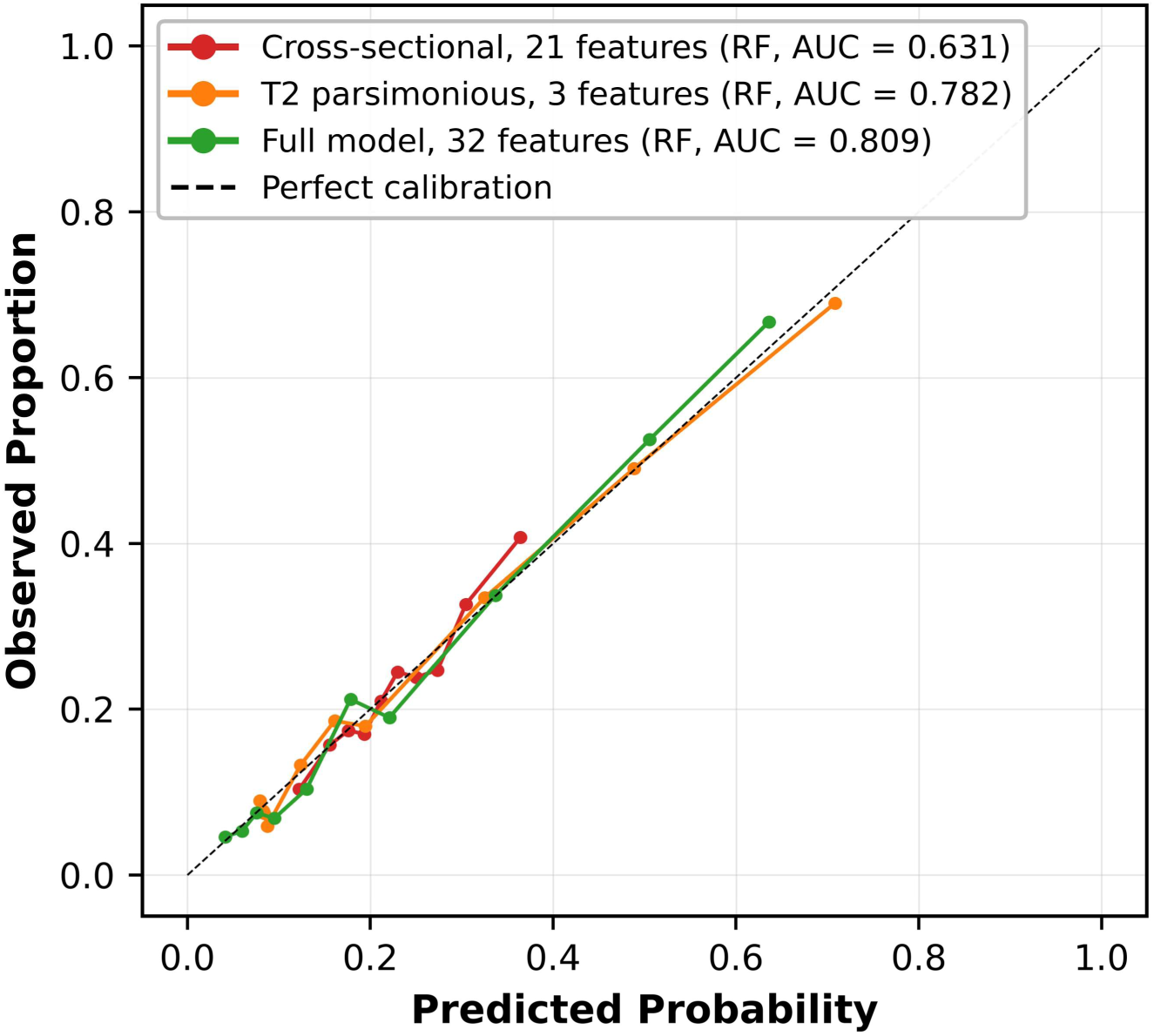
Calibration curves for the three feature sets (random forest).

**Figure S3.**
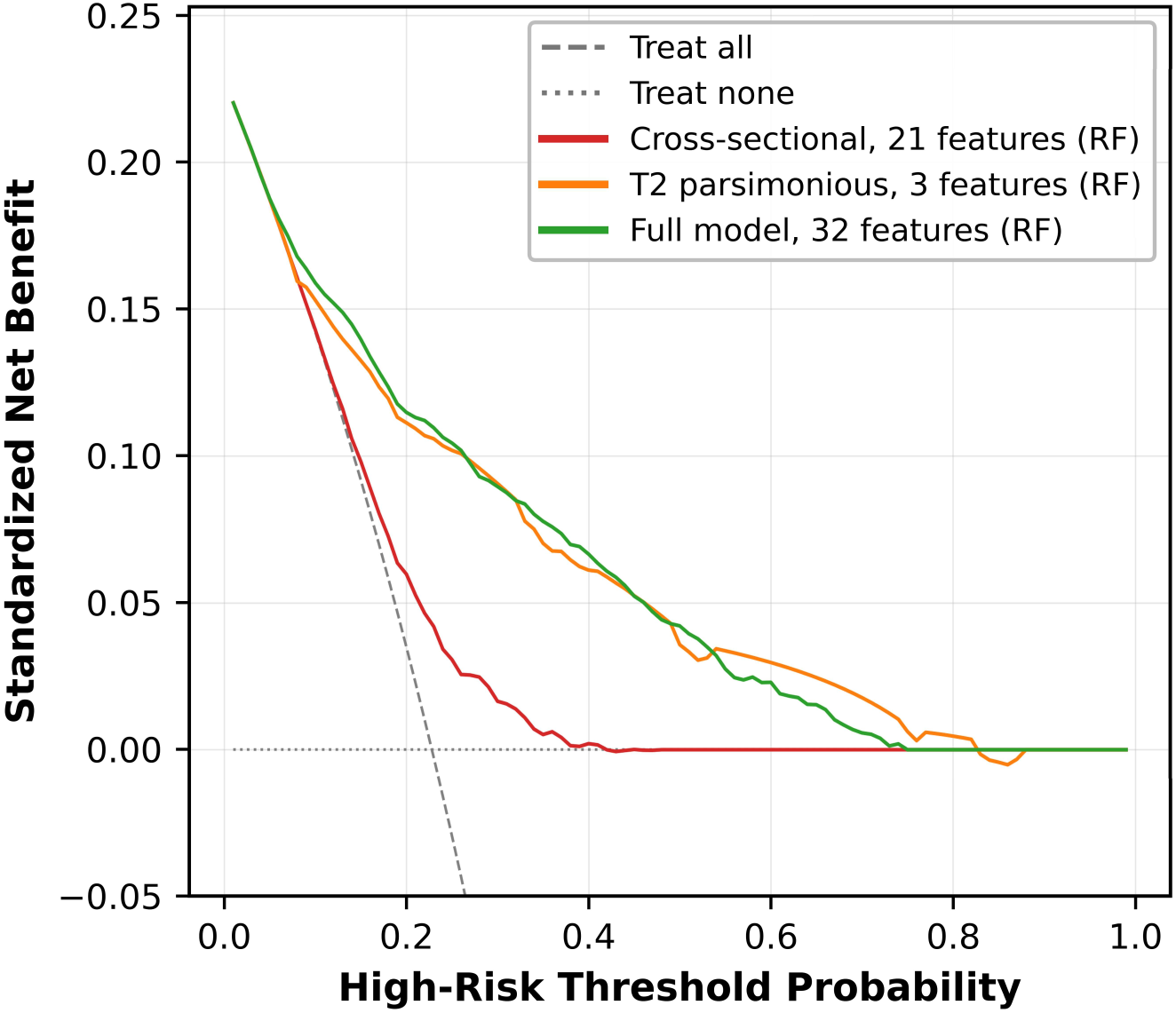
Decision curve analysis (DCA) for the three feature sets.

**Figure S4.**
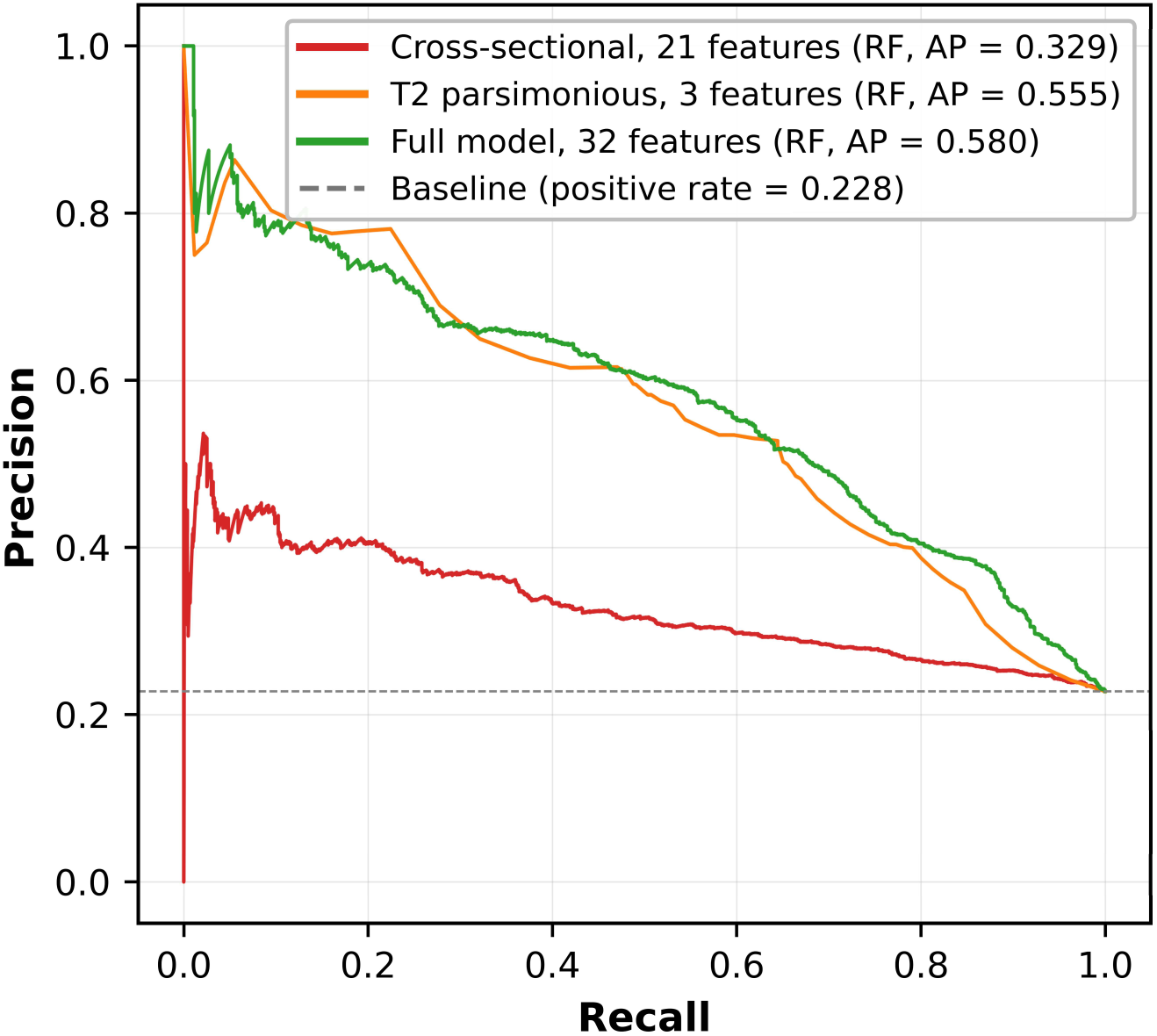
Precision–recall curves for the three feature sets.

**Figure S5.**
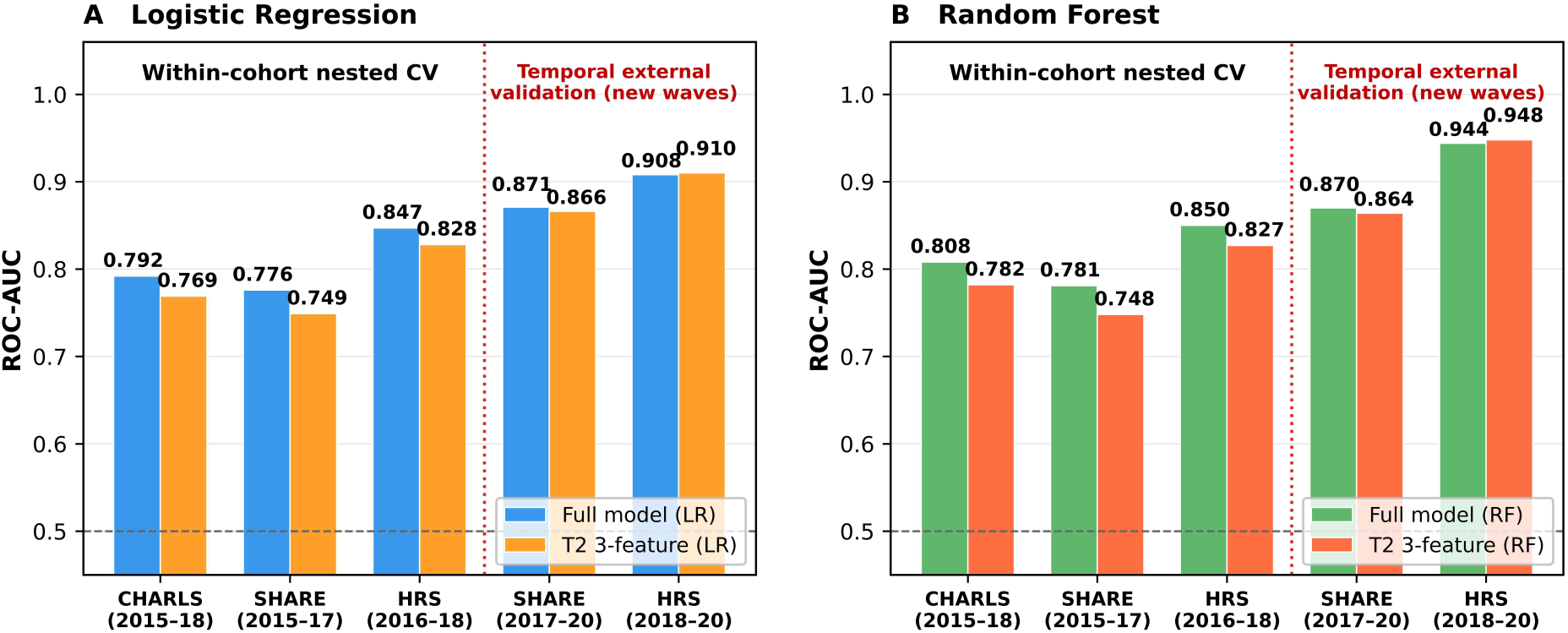
Temporal external validation: models refit on earlier baseline waves and applied to the most recent available wave (SHARE 2017→2020, n=2,851; HRS 2018→2020, n=211, exploratory). LR and RF panels show within-cohort nested cross-validation (T2 and full models) and temporal transport.

